# Development of a simple and highly sensitive virion concentration method to detect SARS-CoV-2 in saliva

**DOI:** 10.1101/2024.04.23.24306243

**Authors:** Yasuko Yamazaki, Uxía Alonso Alonso, Remil L. Galay, Wataru Yamazaki

**Affiliations:** Center for Southeast Asian Studies, Kyoto University, 46 Shimoadachi-cho, Yoshida, Sakyo-ku, Kyoto 606-8501, Japan; Animal Medicine and Health at Institut de Recerca i Tecnologia Agroalimentàries (IRTA) – Centre de Recerca en Sanitat Animal (CReSA), Spain; Department of Veterinary Paraclinical Sciences, College of Veterinary Medicine, University of the Philippines Los Baños, Los Baños, Laguna 4031, Philippines; Kyoto University School of Public Health, Konoe-cho, Yoshida, Sakyo-ku, Kyoto 606-8303, Japan

**Author notes:** Address for correspondence: Wataru Yamazaki, Center for Southeast Asian Studies, Kyoto University, 46 Shimoadachi-cho, Yoshida, Sakyo-ku, Kyoto 606-8501, Japan. (Y. Yamazaki,; U. Alonso Alonso,; R. L. Galay,; W. Yamazaki,).

**Keywords:** Concentration, COVID-19, SARS-CoV-2, Semi Alkaline Proteinase, virion, virus

## Abstract

**Background:** Controlling novel coronavirus pandemic infection (COVID-19) is a global challenge, and highly sensitive testing is essential for effective control. The saliva is a promising sample for high-sensitivity testing because it is easier to collect than nasopharyngeal swab samples and allows large-volume testing.

**Results:** We developed a simple SARS-CoV-2 concentration method from saliva samples that can be completed in less than 60 min. We performed a spike test using 12 ml of saliva samples obtained from healthy volunteer people, and the developed method performance was evaluated by comparison using a combination of automatic nucleic acid extraction followed by RT-qPCR detection. In saliva spike tests using a 10-fold dilution series of SARS-CoV-2, the developed method was consistently 100-fold more sensitive than the conventional method.

**Conclusions:** The developed method can improve the sensitivity of the SARS-CoV-2 test using saliva and speed up and save labor in screening tests by pooling many samples. Furthermore, the developed method has the potential to contribute to the highly sensitive detection of various human and animal viral pathogens from the saliva and various clinical samples.

**Highlight:**

- A method has been developed to detect SARS-CoV-2 from human saliva with 100 times higher sensitivity than conventional methods.
- The developed method combines simple pretreatment within 60 min with conventional nucleic acid extraction and RT-qPCR.
- This method can be applied for more sensitive virus testing from individual saliva.
- This method can potentially be applied to screening more than 100 saliva samples while maintaining the equivalent detection power of conventional methods.
- The method can be adapted to improve the sensitivity of detecting various pathogens from human and animal saliva.

## Introduction

The COVID-19 epidemic continues as of 2023 and remains a public health threat (WHO). Virus detection using purified RNA obtained by extraction kits and real-time reverse transcription quantitative PCR (RT-qPCR) has been universally used due to its high detection sensitivity and low incidence of false negatives due to nonspecific amplification (Lu *et al*., 2020; Vogels *et al*., 2020). Virus detection using saliva has been used to diagnose respiratory infections such as COVID-19 and influenza because of its easy sampling and low burden on patients (Azzi *et al*., 2020; To *et al*., 2017; Vogels *et al*., 2020). However, because of its somewhat lower detection sensitivity compared to nasopharyngeal swab samples, false-negative results occur in samples collected early in infection or late in recovery when viral load is low, meaning that patients who slip through the test may not receive appropriate quarantine measures and become a potential source of infection (Azzi *et al*., 2020; To *et al*., 2017; Vogels *et al*., 2020; Yamazaki *et al*., 2021).

In addition, in the early stages of an epidemic, when there are few positive patients, it is crucial to test a large number of samples from a large number of people for negative confirmation to prevent the spread of infection (Barat *et al*., 2021; Watkins *et al*., 2021). In this case, to save cost and labor, pooling of samples, such as saliva (Barat *et al*., 2021; Watkins *et al*., 2021), and nasal, nasopharyngeal and oropharyngeal swabs (Ayaz *et al*., 2022; Praharaj *et al*., 2020; Pratelli *et al*., 2022) from several people and testing them together is sometimes used for screening.

However, since the positive samples could be diluted by mixing with negative samples, leading to low virus concentration in the pooled sample, the test can become a false negative if it is below the detection limit; hence, accurate detection may not be possible. As a solution, we have developed a method for detecting concentrated viruses in samples through immunomagnetic beads (Yamazaki *et al*., 2019; Makino *et al*., 2020). Still, it is not versatile because it requires specific antibodies for each virus.

The Polyethylene glycol (PEG) precipitation method has been used worldwide to enrich and detect norovirus and other viruses from oysters (Lowther *et al*., 2019; National Institute of Health Sciences, Japan (NIHS) 2010; Yamazaki *et al*., 2022). While this method can concentrate any virus, the presence of sample-derived inhibitors reduces concentration performance (Lowther *et al*., 2019; Miura *et al*., 2018; Yamazaki *et al*., 2022). In our previous studies, we have shown that a combination of a very short, low centrifugation process (900 *g*, 1 min) and a normal centrifugation process (10,000-20,000 *g*, 5 min) as a pretreatment step for simple concentration detection of target bacteria in chicken cecal contents (Sabike *et al*., 2016). Also, we have demonstrated that genetic testing for SARS-CoV-2 is possible without using an extraction kit by digesting human saliva containing potential genetic testing inhibitors with semi-alkaline protease (SAP) (Yamazaki *et al*., 2021). Here, we report the successful development of a new method for the concentration and detection of SARS-CoV-2 from a large volume of saliva by improving and integrating our previously published methods.

## Materials and Methods

### Saliva sampling

Saliva samples were collected from three healthy volunteers, i.e., the three authors of this paper (YY, UAA, and WY), by repeatedly transferring drool collected in the oral cavity into a 50-ml sterile tube. After each saliva was thoroughly mixed by vortexing, the multiple saliva was promptly mixed in a new 50-ml tube to produce approximately 49 ml of pooled saliva. The three saliva samples used were confirmed to be SARS-CoV-2 negative by two RNA extraction methods (conventional and developed) and RT-qPCR detection, as described below, before the experiment.

### Preparation of a 10-fold dilution series of SARS-CoV-2 spiked saliva

A 10-fold dilution series of heat-inactivated SARS-CoV-2 (ATCC VR-1986HK; American Type Culture Collection, Manassas, VA, USA) in PBS was prepared. The pooled saliva was dispensed into four 50-ml tubes of 12.2 ml each. SARS-CoV-2-containing saliva from neat to 10(-4) fold dilutions was prepared by sequentially spiking the 10-fold dilution series of SARS-CoV-2 into the 50 ml tubes containing pooled saliva and then vortexed thoroughly (Table 1).

**Table 1.**
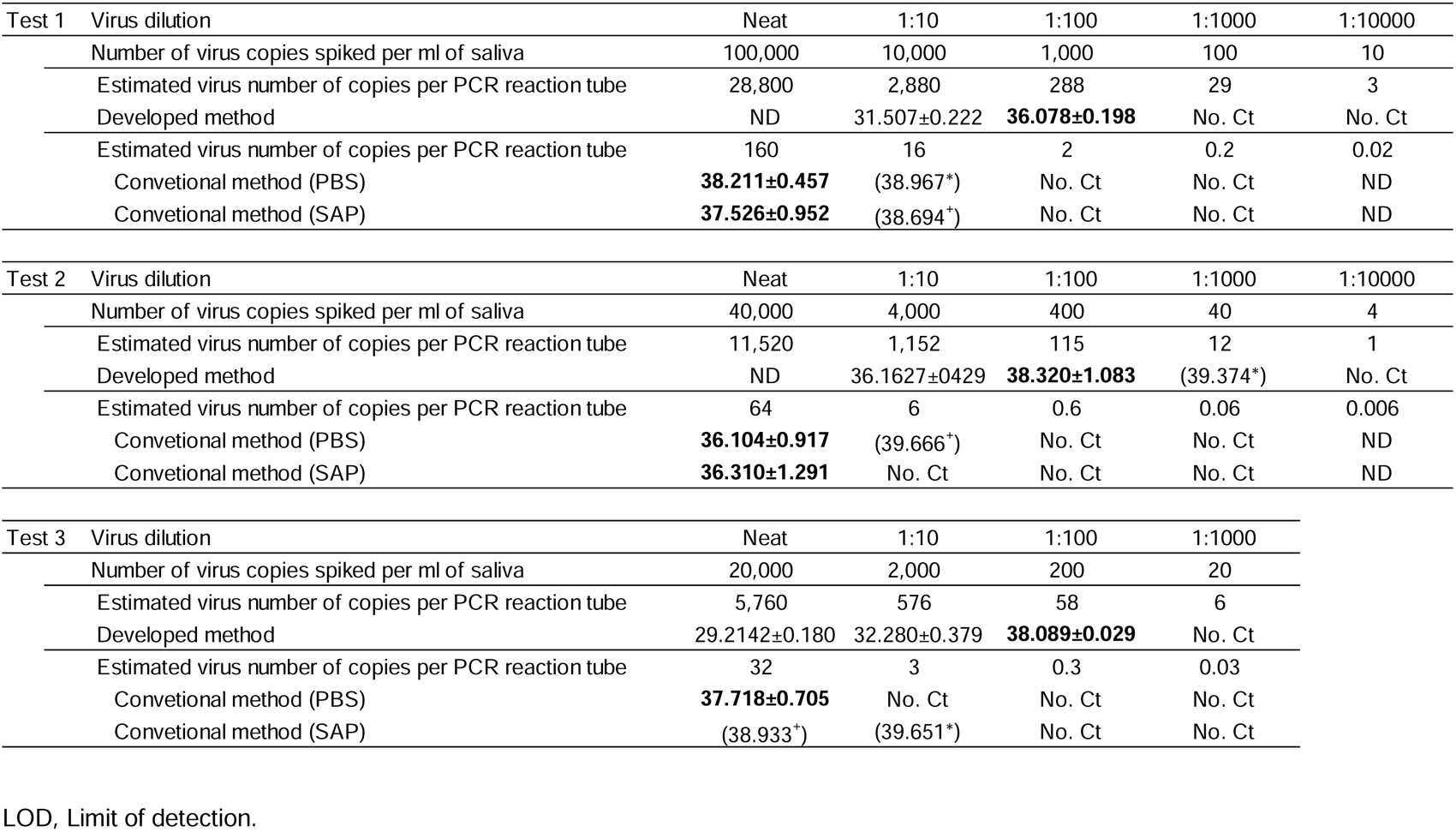
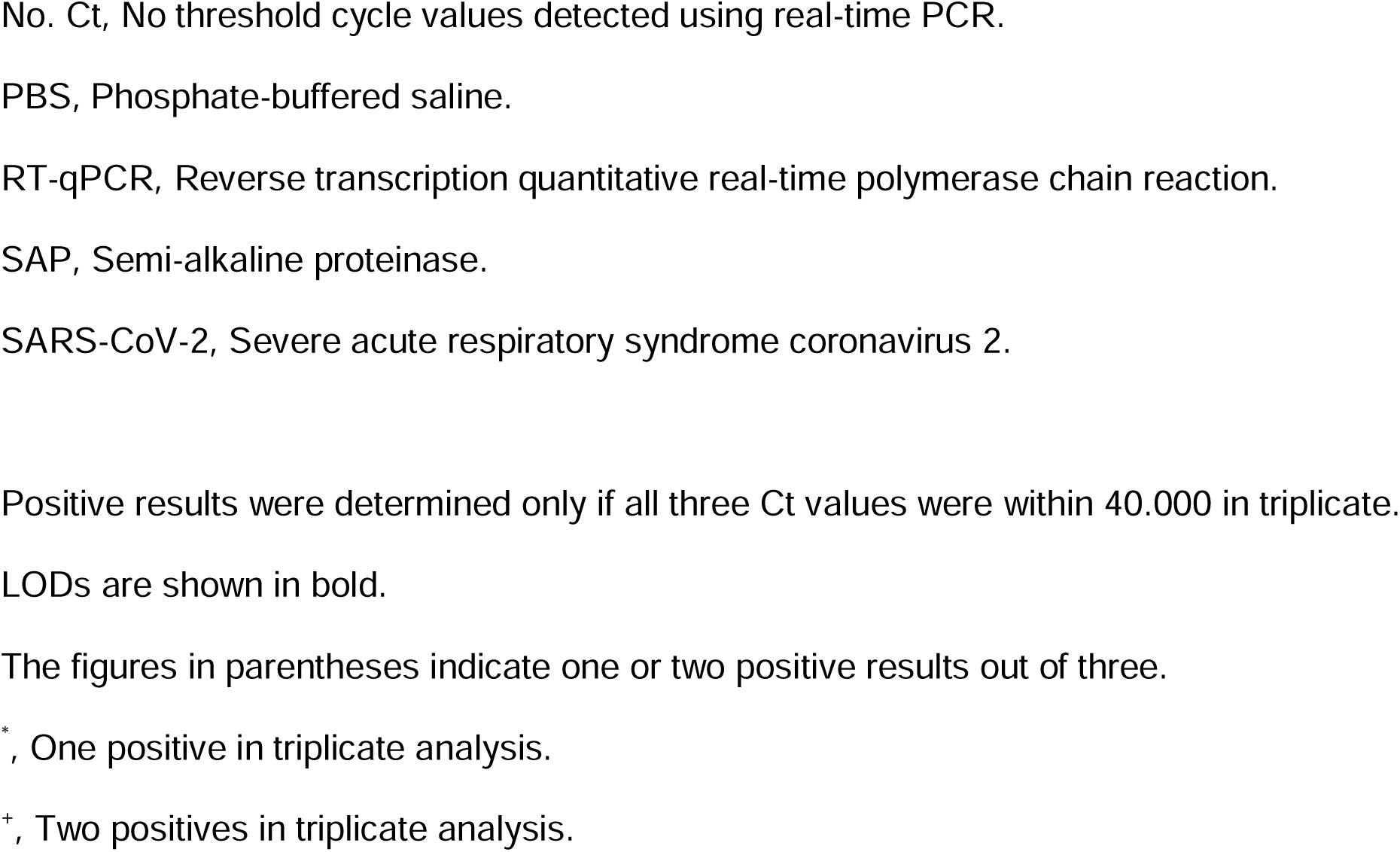
LOD determination of developed and conventional methods using saliva spiked with a 10-fold dilution series of SARS-CoV-2.

### RNA extraction by the conventional method

According to the pathogen detection manual 2019-nCoV issued by the National Institute of Infectious Diseases, Japan (NIID-J), two sets of the 100 μl saliva-spiked SARS-CoV-2 were collected into 1.5-ml microcentrifuge tubes and diluted 1:3 with 300 μl of PBS and sputum homogenizer SAP (Semi-Alkaline Proteinase, Suputazyme; Kyokuto Pharmaceutical Industrial, Tokyo, Japan), respectively. After sufficient vortexing, both (PBS and SAP) of the 1:3 dilutions containing SARS-CoV-2 were centrifuged at 20,000 *g* for 30 min, the former immediately and the latter after 15-min incubation with ten manual inversions mixing every 3 min at room temperature. The resulting 200 μl of the supernatant was transferred in a new 1.5-ml microcentrifuge tube and was set in an automated nucleic acid extractor MagLead 6GC (Precision System Science, Co., Ltd, Matsudo, Japan) with MagDEA Dx SV reagent cartridge (Precision System Science) and were extracted and purified as RNA in 50 μl of distilled water.

### Viral concentration and RNA extraction by developed method

An overview is shown in Figure 1. Specifically, 12 ml of SAP (Kyokuto) was added to the remaining 12 ml of saliva containing SARS-CoV-2. After vortexing, the mixture was kept at room temperature for 15 min. During the 15-min incubation, ten inversion mixings were performed manually every 3 min. Then, 4,000 *g*, 5 min initial centrifugation was performed. Taking care not to inhale the pellet derived from the formed saliva components, 18 ml (75% of the initial mixture volume) of the centrifugal supernatant was prudently transferred to a new 50-ml tube. 15 ml of SAP (Kyokuto) was added and incubated for 15 min at room temperature. Then, 13.2 ml of PEG solution (40% PEG-NaCl, see details in our previous publication, Yamazaki *et al*., 2022) was added and thoroughly mixed by vortexing, followed immediately by a second centrifugation at 8,000 *g* for 20 min. The supernatant was carefully removed after the second centrifugation. To prevent contamination, 100 μl of PBS in a 1-ml long tip, which is longer than the 50 ml tube, was added. Pipetting was performed with the 1-ml long tip set pipet from the bottom of the tube to the area where the pellet had adhered during the first centrifugation, where precipitates of PEG-virus particle complex are assumed to be attached, although it cannot be seen with the naked eye. In addition, to completely detach any PEG-virus particle complex precipitates that may still be adhering to the tube wall, the 1-ml short tip was added to the 50-ml tube, the lid was closed, and the tube was thoroughly vortexed. Approximately 200 μl of the mixture of about 100 μl of PBS (containing PEG-virus particles) added to the around 100 μl of supernatant remaining on the inner wall of the 50-ml tube obtained by flushing was transferred into a 1.5-ml screw cap tube using a 1-ml long tip. The mixture was then extracted and purified as 50 μl of RNA using an automated nucleic acid extractor (Precision System Science), as described above. When the extracted RNA could not be tested immediately, it was stored at - 80°C until use.

**Figure 1.**
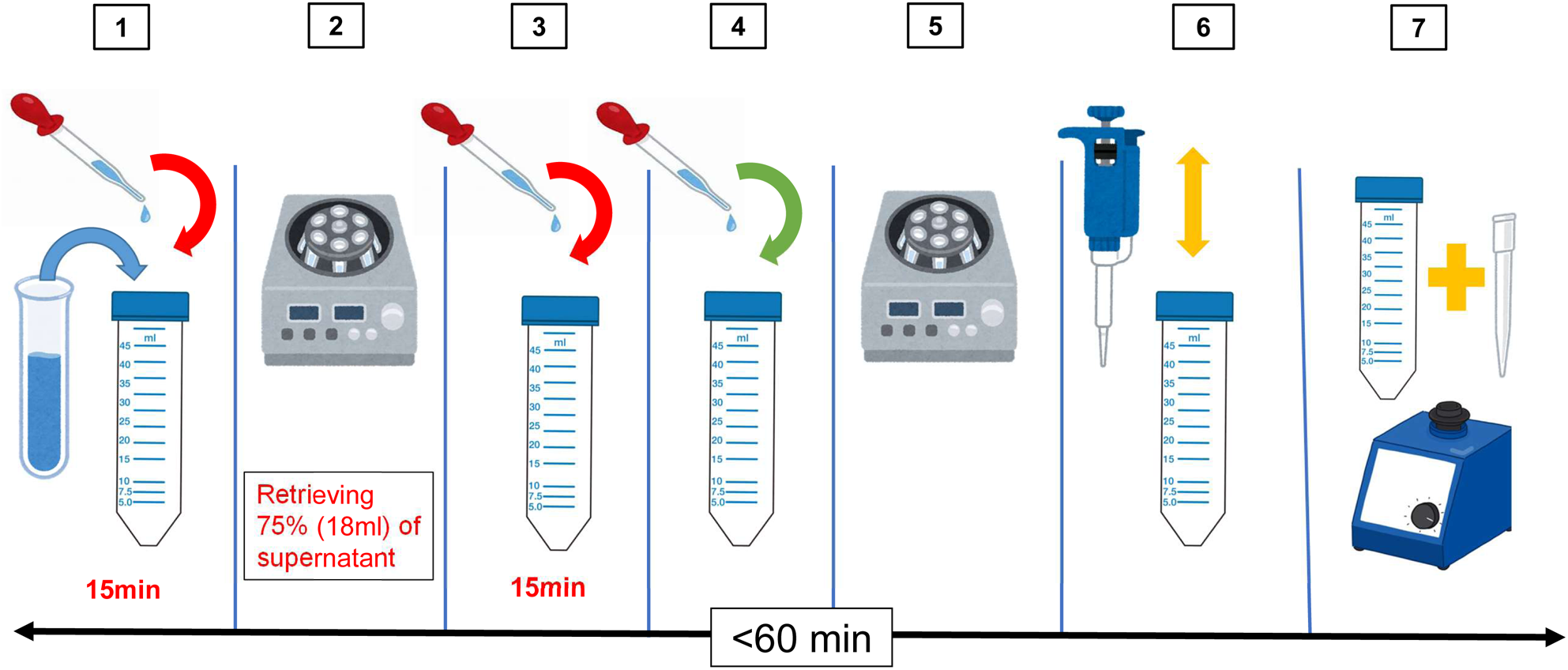
Developed protocol for virion concentration from saliva. SAP, Semi-alkaline proteinase. 1. Mix 12 ml of saliva with 12 ml of SAP (1:1) and keep for 15 min at room temperature. 2. After centrifugation at 4,000 *g* for 5 min, carefully transfer 18 ml of the supernatant into a new 50-ml tube. 3. Add 15 ml of SAP to 18 ml of the supernatant, mix using a vortex and then keep at room temperature for 15 min. 4. After adding 13.2 ml of PEG-NaCl solution, mix by vortexing. 5. After centrifugation at 8,000 *g* for 20 min, carefully discard the supernatant. 6. Add 100 μl of PBS and dissolve the invisible precipitates by pipetting and scraping with a 1-ml long tip (10 times each). 7. Place a 1-ml short tip into a 50-ml tube and vortex to completely dissolve the precipitate (supernatant residue after flushing + PBS ≒ 200 μl), and then transfer to an RNA extraction tube.

### Conducting RT-qPCR and determination of LOD

RT-qPCR was performed with 4 μl of the extracted RNA in 20 μl of the reaction mixture using a QuantStudio 3 (Thermo Fisher Scientific, Inc., Waltham, MA, U.S.A.), according to the method by Lu and colleagues (2020). The amplification time was slightly extended to ensure detection, as described below. Details of the reagents used are as follows: 20-µl RT-qPCR reactions comprised 10 µl of SuperScript III Platinum One-step RT-qPCR 2x reaction (Thermo Fisher Scientific), 0.4 µl of SuperScript III/Platinum *Taq* Mix (Thermo Fisher Scientific), 2 µl of primer (Hokkaido System Science Co. Ltd., Sapporo, Japan) probe (Integrated DNA Technologies, Inc, Singapore) mix for SARS-CoV-2 N2 detection reported by Emergency Use Authorization issued by the US Food and Drug Administration issued by the FDA (Lu *et al.,* 2020), 2 µl of magnesium sulfate (50 mM, Thermo Fisher Scientific), 0.2 µl of Rox Dye (Thermo Fisher Scientific) diluted 1:5 with distilled water, 1.4 µl of nuclease-free water, and 4 µl of the RNA template. The cycling conditions were as follows: one cycle at 50°C for 900 sec and 95°C for 120 sec, followed by 50 cycles each at 95°C for 15 sec and 55°C for 60 sec. The automatically calculated Ct value was adopted, and the Ct cut-off value was set at 40.000. Positive results were determined if all three Ct values were within 40.000 in triplicate. Samples that showed only one or two positive Ct values in the triplicate analysis were interpreted as negative. The mean and standard deviation of the Ct were calculated for all samples interpreted as positive.

## Results

Tables 1 and 2 show that the developed concentration method enabled 100-fold more sensitive detection than the conventional method by adding only a simple pretreatment within 60 min before the conventional extraction method. The developed method required only a centrifuge machine for 50 ml tubes up to 8,000 *g* and a vortex for mixing the liquid in the 50 ml tubes and did not need expensive equipment such as an ultracentrifuge.

**Table 2.** Comparison of LOD between developed and conventional methods by RT-qPCR.

|  | LOD per ml of saliva | LOD per RT-qPCR<br>reaction tube |
| --- | --- | --- |
| Developed | 200 | 58 |
| Conventional (PBS) | 20,000 | 32 |
| Conventional (SAP) | 40,000 | 64 |

LOD, Limit of detection.
PBS, Phosphate-buffered saline.
RT-qPCR, Reverse transcription quantitative real-time polymerase chain reaction.
SAP, Semi-alkaline proteinase.

As shown in Tables 1 and 2, the conventional method required 20,000 to 40,000 copies of virus per ml of saliva to detect SARS-CoV-2, whereas the developed method required only 200 copies of virus per ml of saliva. In other words, the developed method was at least 100 times more sensitive than the conventional method. Furthermore, a comparison of the number of viral copies per RT-qPCR reaction tube showed that 32 to 64 and 58 copies were required for detection by the conventional and developed methods, respectively. Namely, LOD per reaction tube was comparable for the two methods.

## Discussion

In Japan, the airport quarantine for COVID-19 recommends collecting and submitting approximately 5 ml of saliva. Still, following the protocol of the NIID-J (2020), 200 μl of the supernatant is generally used after diluting 200 μl within the range of 1:1 to 1:3 ratios and centrifuging at 20,000 *g* for 30 min. In other words, only 50-100 μl of saliva is used for the testing, and the remaining saliva of around 5 ml is used for nothing but retests. If all the unused samples were simply submitted to the concentration method developed in this study, the detection sensitivity would be dramatically increased up to 100 times. Hence, a more accurate quarantine control measure would be possible.

The disadvantage of pool testing is decreased LOD (Barat *et al*., 2021; Praharaj *et al*., 2020; To *et al*., 2017; Watkins *et al*., 2021), but the developed concentration method can solve this problem. Hence, the developed method is ideal for labor-saving large-scale screening in the early stages of an outbreak when the positivity rate is low. When a new variant emerges in the future, the concentration method could be used as a large-scale screening test to reasonably enhance quarantine control measures and contribute to efficient epidemic control. The developed method is theoretically capable of 180-fold virus concentration (Figures 1 and 2). Since the actual measured value is about 100-fold (Tables 1 and 2), it should be noted that the recovery rate could be reduced by approximately 50%. In the conventional method, the 12-ml saliva sample is equivalent to 240 pooled samples of 50 µl of saliva per individual. Still, considering the recovery rate, a pool of more than 100 samples can be expected to have a detection sensitivity equivalent to or better than that of the conventional method.

**Figure 2.**
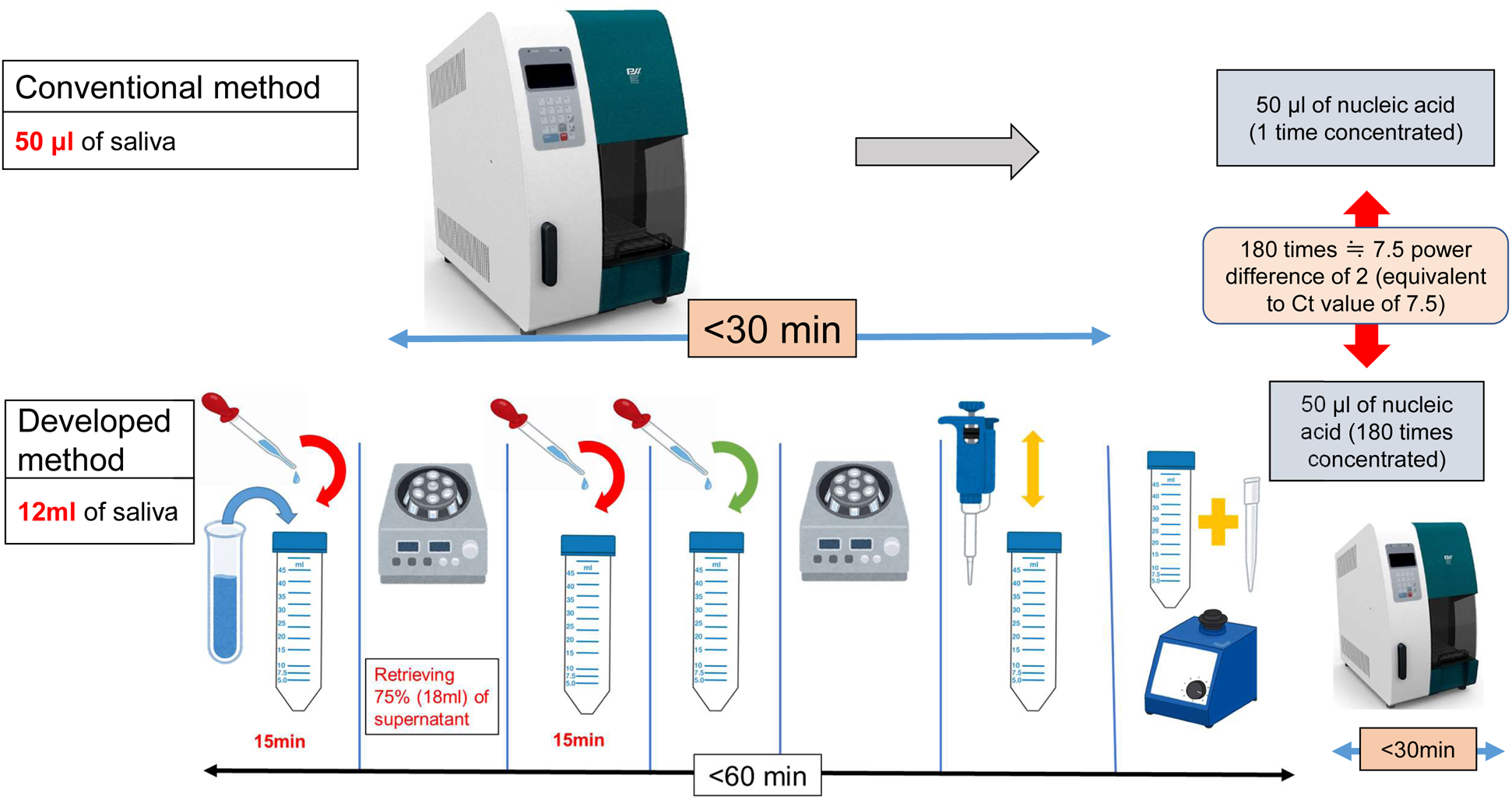
Virus detection from saliva with developed and conventional methods. SAP, Semi-alkaline proteinase. Conventional method: Nucleic acid extraction of 200 μl of centrifuged supernatant comprising of saliva 50 μl + PBS or SAP 150uL (4x dilution). Developed method: Nucleic acid extraction after a simple concentration of 12ml of saliva. The illustrations are cited from the following sources, all used in compliance with the terms and conditions. Pipettes and Micropipettes: Irasutoya (irasutoya.com) Vortex mixer: Kagaku Irasuto (science-illust.com) 50 ml tubes: Kenkyu Net (wdb.com/kenq/illust) Automatic nucleic acid extractor: Precision System Science Co. Ltd. (pss.co.jp/)

In our previous study, an immunomagnetic bead method using specific antibodies was successfully used to detect influenza A viruses added to PBS, duck feces, and chicken meat at 10- to 1,000-fold sensitive concentration (Yamazaki *et al*., 2019; Makino *et al*., 2020). Although this method is extremely sensitive, it is not very versatile because it requires the preparation of specific antibodies for each virus species and has the disadvantage that the LOD is reduced when the samples contain many inhibitory substances, such as components of the duck feces and chicken meat. In the present study, this problem has been successfully overcome by improving the pretreatment method with the combination of SAP, a sputum dissolving agent, and the PEG precipitation method to achieve highly sensitive SARS-CoV-2 concentration and detection in saliva.

The PEG precipitation method customarily includes an overnight 4°C incubation process for virion capture (Lowther *et al*., 2017; Yamazaki *et al*., 2022). However, the NIHS protocol (2010) states that the virion-PEG complex can be recovered from the oyster midgut gland immediately by centrifugation at 8,000 *g* for 20 min without an overnight incubation. In the present study, we referred to this finding and confirmed that SARS-CoV-2 in saliva could be concentrated immediately after being centrifuged at 8,000 *g* for 20 min without needing overnight incubation before the centrifuge, as expected. This allowed us to establish a rapid protocol successfully. This study’s limitations include the inability to evaluate SARS-CoV-2-positive clinical samples and the fact that, although the concentration process is simple, the number of steps involved requires care by the examiner to avoid contamination and laboratory infection.

When the PEG-NaCl solution is mixed with a liquid sample containing trace amounts of virions, PEG, a polymer, adsorbs water molecules, causing the virions to aggregate. By centrifuging, the agglomerated virions-PEG complex can be precipitated on the wall of the tube. On the other hand, if the liquid sample contains impurities, this reaction is inhibited, and the recovery rate of virus particles is reduced. As shown in Table 1, the developed method is more sensitive than the conventional method, but the estimated number of copies of virus per reaction tube may be higher for the developed method to obtain a positive result. For example, as shown in Tables 1 and 2, Test 2 showed Ct values at 36.104 and 36.310 for the conventional method (64 copies/RT-qPCR reaction tube) versus the Ct value at 38.320 for developed method (115 copies/RT-qPCR reaction tube). This suggests that, although carefully collected, there is some residual material in the supernatant after centrifugation that inhibits virion-PEG complex formation or that there reduced viral recovery, resulting in an increase of the Ct value in the developed method.

In the developed method, only 75% of the saliva centrifugal supernatant is used and the remaining 25% must be discarded without use, as shown in Figure 1, Process 2. In our preliminary experiment, the maximum amount of centrifugal supernatant corresponding to more than 90% was collected, and concentration detection was attempted. However, contrary to expectations, the recovery rate was more than 10 times lower than the theoretical value (data not shown). In this case, a large pellet was identified on the tube by the naked eye in process 6 of Figure 1. In other words, although the centrifugal supernatant appeared clear by the naked eye observation in Processes 2-5, we speculated that saliva components were mixed in and inhibited the formation of virion-PEG complexes. Therefore, we did not use the lower portion of the centrifugal supernatant, which was presumed to contain more saliva components due to the gradient caused by centrifugation but used only the upper 75% to obtain a stable recovery rate.

Nevertheless, the developed method has the potential to solve the technical limitation of conventional genetic testing methods, i.e., the problem that samples carrying trace amounts of the virus have been judged as false negative because they are below the LOD (Barat *et al*., 2021; Praharaj *et al*., 2020; To *et al*., 2017; Watkins *et al*., 2021; Yamazaki *et al*. 2019). For example, rabies is transmitted from dogs to humans via dog saliva. Still, definitive diagnosis requires dog brain emulsion containing large amounts of rabies virus, not the dog saliva, due to lack of the virus amount, and there is an animal welfare issue of euthanasia of dogs for sampling. The oral pulse oximeter used for anesthesia monitoring in veterinary clinics caused the transmission of severe fever thrombocytopenia syndrome (SFTS) virus transmission to cats due to contamination. However, the low amount of the virus was not detectable (Mekata *et al*., 2023). Both cases demonstrate the problem that current genetic testing cannot accurately detect trace amounts of virus in saliva, leading to false-negative diagnoses.

## Conclusions

We have successfully developed a simple and highly sensitive method for concentrating SARS-CoV-2 in saliva. We demonstrated that the developed method has an extremely sensitive detection performance that is at least 100 times higher than the conventional method. We further showed that this method has the potential to screen more than 100 saliva samples with power comparable to conventional extraction methods. In the future, this method may be applied to highly sensitive diagnosis of various human and animal viral infections that can be tested from saliva, as well as to rapid screening by pooled testing of a large number of samples.

## Ethics approval

The Ethics Committee of Kyoto University Graduate School and the Faculty of Medicine approved this study (R2379), and consent to participate was waived from three volunteer saliva sample donors who are also authors.

## Declaration of Availability of data and materials

All data obtained in this study is included in the paper and Supplemental Table 1. In addition, the data sets in this study are available from the corresponding author upon reasonable request.

## Declaration of Consent for publication

Consent for publication has waived the need to obtain informed consent from three volunteer saliva sample donors who are also authors.

## Declaration of Generative AI and AI-assisted technologies in the writing process

During the preparation of this work, the authors used DeepL (DeepL SE, Cologne, Germany) to improve readability and language. After using this tool, the authors reviewed and edited the content as needed and took full responsibility for the content of the publication.

## Declaration of Submission

The authors confirm that this manuscript or data has not been previously published and is not being considered for publication elsewhere. The authors further confirm that all authors have contributed to the study and have approved the final version.

## Authors’ contributions

**YY**: Data curation, Formal analysis, Funding acquisition, Methodology, Resources, Validation, Visualization, Writing - original draft, Writing - review & editing. **UAA**: Methodology, Resources. **RLG:** Formal analysis, Writing - review & editing. **WY**: Conceptualization, Data curation, Funding acquisition, Methodology, Project administration, Resources, Supervision, Validation, Visualization, Writing - original draft, Writing - review & editing.

## Abbreviations

ATCC: American Type Culture Collection
LOD: Limit of detection
PBS: Phosphate-buffered saline
PEG: Polyethylene glycol
RT-qPCR: Reverse transcription quantitative real-time polymerase chain reaction
SAP: Semi-alkaline proteinase
SARS-CoV-2: Severe acute respiratory syndrome coronavirus 2

## Data Availability

All data produced in the present study are available upon reasonable request to the authors.

## Data Availability

All data produced in the present study are available upon reasonable request to the authors.

